# Fecal sample biobanking for breast cancer research focused on the gut microbiome

**DOI:** 10.1101/2025.10.31.25339256

**Authors:** Lusine Yaghjyan, Neha Goel, John Heine, Doratha A. Byrd, Sugriva E. Forsyth, Erin Fowler, Kathleen M. Egan

**Affiliations:** University of Florida, College of Public Health and Health Professions and College of Medicine, Department of Epidemiology, Gainesville, FL, USA; Department of Surgery, Division of Surgical Oncology, School of Medicine, University of Miami Miller, Miami, FL; Sylvester Comprehensive Cancer Center, School of Medicine, University of Miami Miller, Miami, FL; Moffitt Cancer Center, FL, USA

**Keywords:** gut microbiome, breast cancer, mammographic breast density, breast cancer risk

## Abstract

**Background:** Gut microbiome is an emerging potentially modifiable contributor to breast health, including breast cancer (BCa). To advance prevention research in this area, we established a prospective biobanking cohort of cancer-free women.

**Methods:** Eligible women were ≥40 years old, had no cancer history and no recent antibiotic use. Women were enrolled during screening mammography visits at three imaging centers in Florida (February 2021-June 2024), completed a BCa risk factor survey, and underwent body measurements. We collected digital mammograms and stool/urine/saliva samples. Optionally, women completed NIH’s Diet History Questionnaire and a neighborhood stress questionnaire. Mammographic breast density (MBD) was assessed using established computerized approaches.

**Results:** We recruited 733 cancer-free women (49% Caucasian, 21% African American, 26% Hispanic, and 4% from mixed/other races). The average age was 60 years (range 40-92); the majority (68.3%) were postmenopausal. BCa risk factor, neighborhood stress and diet questionnaires were completed by 97%, 65% and 58% of participants, respectively. Urine, saliva, and mammograms were available for all women; 83% also returned stool samples.

**Conclusions:** We have established a representative cohort of screen-aged women with comprehensive BCa risk factor data, biospecimen collection, and MBD.

**Impact:** This unique resource provides opportunity for future gut microbiome-focused BCa prevention research.

## Introduction

Breast cancer (BCa) remains by far the leading cancer diagnosis and the second leading cause of cancer mortality. While BCa treatments have improved in recent years [1] early detection and prevention remain key to reducing BCa incidence and the burden of disease. Several recent studies suggest that gut microbiome may represent an emerging potentially modifiable contributor to a woman’s risk for breast cancer.

The populations of microbes that inhabit human bodies have complex relationships with their hosts. Microbes inhabiting the gut mediate key metabolic, physiological and immune functions [2-5] and perturbations in these functions influence health and disease [4-10].

The intestinal microbiome contributes to important protective, metabolic, and structural functions relevant to breast cancer, including host immunity; metabolism of a wide range of compounds (steroid hormones, phytochemicals, nitrates, and xenobiotics); integrity of the epithelial barrier and absorption; and host energy balance and nutritional status [11-14]. As a result, the dynamic population of intestinal microbes can influence metabolism and absorption of compounds with carcinogenic properties, modify levels of endogenous factors implicated in breast carcinogenesis (for example, steroid hormones), and alter inflammation and host susceptibility to oncogenic factors [15]. In particular, variations in the gut microbiome’s genes that are capable of metabolizing estrogens could affect risk of estrogen-driven breast cancer and offer future gut microbiome-directed targeted interventions to reduce breast cancer risk [16-18].

In spite of compelling theoretical grounds for a role of the gut microbiome in breast cancer, epidemiological evidence is limited due in part to the exceptional challenges of studying this exposure in observational research. In retrospective case-control studies, gut microbiome profiles in cancer cases may reflect changes in microbial composition that occur as a result of the cancer diagnosis or treatment, with potential for reverse causality [19]. In prospective cohort studies, investigations of the gut microbiome are impractical in the near term due to the large numbers of subjects that must be enrolled, long follow-up, and repeat sampling that may be required to capture change in gut microbiome profiles over many years of follow up. An alternative strategy to avoid these challenges is to study relationships of the gut microbiome with two of the leading, well-documented risk factors for breast cancer, i.e. mammographic breast density and endogenous hormone profiles. Importantly, use of well-known intermediate phenotypes has been recognized as an efficient alternative for studying chronic diseases that could also provide valuable information for primary cancer prevention [20-24].

To advance knowledge in this understudied area, we established an infrastructure that allowed us to prospectively recruit cancer-free women undergoing routine mammographic screening for breast cancer in the State of Florida with collection of urine, stool, and saliva samples and comprehensive data on critical breast cancer risk factors. The Florida population is among the most diverse in the nation and includes a large segment of traditionally underrepresented persons in research, including those of African American/Hispanic race, and lower socioeconomic status with poor health care, offering unprecedented opportunity for expansion of research into these populations. This unique infrastructure represents an outstanding resource for research on the gut microbiome and implementation of novel integrative molecular epidemiology approaches to the studyof breast cancer etiology and disparities.

## Materials and Methods

### Study population and recruitment

Recruitment took place between February 2021 and June 2024. Participants were enrolled at imaging centers affiliated with three Florida-based institutions: University of Florida (UF) in Jacksonville; Moffitt Cancer Center (MCC) in Tampa; and the University of Miami (UM). Collaborating centers are located in north, central and south Florida, respectively, and offer broad coverage of the state’s nearly 23 million residents. UF in Jacksonville serves the local community that comprises the state’s largest population of African American women. UM Sylvester Comprehensive Cancer Center, a NIH-designated cancer center serves the greater Miami area, along with south and central America, and is home to the state’s largest Hispanic community. MCC, a NIH-designated Comprehensive Cancer Center, serves a 15-county region on Tampa Bay in west central Florida comprised of more than 6 million persons (29% of the state’s population).

At MCC, participants were identified through the IRB-approved protocol that provides infrastructure for studies of lifestyle and genetic determinants of breast health measures in screening-aged women. Ethnic composition of MCC clinic patients mirrors that of the underlying population in Moffitt’s catchment area (∼70% Caucasian, 15% Hispanic, and 15% African American women). At UF Jacksonville, participants were identified through the UF Jacksonville Imaging Centers. The Centers are accredited by the American College of Radiology and provide state-of-the-art screening and diagnostic mammography. Jacksonville is the largest (excluding the large metro areas of Miami and Tampa) and fastest-growing city in the state of Florida. Located in Duval County, Jacksonville has the state’s largest African American population. Ethnic composition of Jacksonville clinic patients undergoing screening mammography is similar to the distribution in the catchment area (50% African American, 40% Caucasian, and the remaining ∼10% a mix of Asian, Native American, and other races). At UM, participants were identified through the Posner Breast Imaging Center at the NCI-designated Sylvester Comprehensive Cancer Center (SCCC). Miami is home to a diverse population with high concentrations of native and immigrant White and Black Hispanics from the Caribbean and Latin America. Racial composition of Miami clinic patients undergoing screening mammography is similar to the distribution in the catchment area (∼20% Caucasian, ∼70% Hispanic; and ∼10% African American).

Women were recruited by study coordinators dedicated to this project in clinics during scheduled mammography appointments. IRB authorization for the project was maintained separately at each of the 3 project sites. Written informed consent was obtained from all study participants. Because of the infrastructure building nature of the project and implied access of resources to the research community throughout Florida and beyond, the subjects were made aware that 1) unnamed researchers would have access to their data and biospecimens in the future, 2) resources will support testing of currently unspecified hypotheses, 3) resources will be maintained indefinitely, 4) future research may involve genetic testing of samples and 4) no test results will be made available to participants, in addition to statements on the standard risk and benefits that apply in observational research studies with scope of the planned resource-building project. A $20 gift card was offered to women in appreciation of their time and effort upon completion of the in-clinic portion of the study. An additional $20 gift card was sent to the participant upon receipt of the stool sample.

Women were eligible for this study if they had no cancer history (other than non-melanoma skin). Further exclusions included: i) any oral/IV antibiotics within 30 days and/or, ii) more than two separate antibiotic regimens within the previous 3 months.

### Data collection

Women were approached by study coordinators in study clinics at MCC, UF and UM, given an explanation of the study, and offered an opportunity to enroll. In addition, at UF we distributed information about the study via other primary care prevention offices affiliated with the UF Jacksonville and women interested in participation were then able to set up an appointment with the study coordinator for their participation. At UM, women had the option of completing study procedures in clinic or in their homes during scheduled in-home visits by study coordinators. Eligible women provided consent and completed an electronic survey covering demographics, established breast cancer risk factors (comprehensive reproductive history, including menopausal hormone use, detailed family history of breast cancer, alcohol consumption, and smoking); menopausal symptoms; medications (including recent use of antibiotics); quality of life and other psychosocial factors; and breast screening and medical history). A link to the electronic survey was emailed to women wishing to complete the survey at home. Additionally, women were offered with an opportunity to complete two questionnaires capturing diet and residential history. Information on usual diet in the preceding 12 months was collected via the Diet History Questionnaire food frequency questionnaire (DHQII) developed and administered through the National Cancer Institute that assesses diet and supplement use including probiotics during the previous year and produces correlations for energy, macronutrients, and several vitamins and minerals, consistent with other widely used FFQs [25]. DHQII has been successfully validated in previous studies [25,26]. Women agreeing to complete diet assessment were sent a link to the DHQII that includes an embedded unique sequence of digits that allows each subject’s data to be identified. Diet data is processed with Diet*Calc software developed by the NCI to generate specific nutrient and food group intake estimates. The data is maintained by the NCI and will be available for download for diet-related investigations. The Neighborhood Social Environment Adversity Survey captured information on neighborhood stress [27].

Standard body composition measurements were taken during the clinic or at-home (for UM) visit. For each participant, we recorded height and weight, circumference of the waist and hips (to calculate a waist-to-hip ratio), and chest circumference. Vital signs (pulse and blood pressure) were also recorded for approximately half of the participants. All consents and data were collected and managed using centralized RedCap data management system developed and maintained by the Participant Research, Interventions, and Measurement Core at MCC.

### Sample collection

A clean-catch spot urine specimen was collected at recruitment in study clinics. Samples were aliquoted and stored at −80ºC according to standard operating procedures at each institution, 90% within approximately one hour of collection. Times of urine collection and freezer storage were recorded for all urine samples. The samples from UF and UM were periodically shipped to MCC for centralized banking. Salivary samples were collected via ‘Oragene’ kits as a source of germline DNA and shipped to MCC for long-term banking.

Subjects were provided with a stool collection kit with detailed instructions, a stool collection questionnaire, and postage paid mailing kits for return of the materials according to methods that are used successfully in ongoing funded research projects and pilot investigations [28,29]. Stool samples and stool collection questionnaires were returned by the participants directly to MCC where they were immediately stored in −80^°^C freezer. Times of sample collection and receipt by the Tissue Core at MCC were recorded for all stool samples. The median time between stool sample collection and receipt by MCC was 3 days (range 0-6) for samples returned via FedEX shipment; in a subset of women (74), most enrolled at UM or UF, shipment was delayed by a week or more after collection of the sample for a variety of circumstances

Women also provided consent for release of pathology results and tissue samples in the event of breast biopsy. Formalin-fixed histologic sections will be requested from pathology labs and stored at MCC as they become available.

### Mammogram collection and mammographic breast density assessment

Full field digital mammograms were obtained for all women. All three clinics have Hologic Selenia 3-dimensional (3D) Digital Breast Tomosynthesis units. UF-Jax and UM transferred zipped (compressed) digital image data to MCC using a secure cloud content management system. Upon receipt, DICOM images from all sites were inspected for quality and stored on a dedicated secure server at MCC. Anonymization procedures were performed to remove PHI from stored image data. As the primary measure, percent breast density was determined using methods developed by Heine et al. [30-33] that compute percent of the breast occupied by the dense (epithelial/stromal) tissue relative to the total breast area [30,34]. PD is regarded as the *de facto* standard for MBD determination [35] as it has been shown repeatedly to produce a validated measure of risk regardless of the mammographic method [36-39]. Because breast density of the right and left breast for a woman are strongly correlated [40], the average density of both breasts was used as the final mammographic breast density measure for each woman.

### Data Sharing

The authors recognize and support the principles of resource sharing that have been endorsed by the NIH. Access to data and biospecimens (urine, residual stool sample and germline DNA) from the study described in this report is extremely valuable for future research. Therefore, investigators are encouraged to apply for use of these resources. Access decisions will be based on the following general principles: 1) Scientific merit, 2) Overlap with other approved protocols, and 3) Submission of a research plan appropriate to answer the study question(s). Review of applications and final decisions will be made by a steering committee comprised of the overall project PIs (Drs. Egan, Yaghjyan, and Goel) and other members of the research team. The committee will consider scientific merit, ethical and legal factors, and any potential conflicts of interest on the part of the applicant, with fair and transparent deliberation. Upon acceptance, de-identified samples and/or data will be released with Committee approval, and after establishing a Data Sharing or Material Transfer Agreement between institutions. The investigators have not budgeted costs for distribution of resources to end users. Hence, costs for data downloads or retrieval/ shipment of samples will be borne by researchers with approved protocols making the request. Further information can be obtained from the corresponding author.

### Data availability

The authors recognize and support the principles of resource sharing that have been endorsed by the NIH. Access to data and biospecimens (urine, residual stool sample and germline DNA) from the study described in this report is extremely valuable for future research. Therefore, investigators are encouraged to apply for use of these resources. Access decisions will be based on the following general principles: 1) Scientific merit, 2) Overlap with other approved protocols, and 3) Submission of a research plan appropriate to answer the study question(s). Review of applications and final decisions will be made by a steering committee comprised of the overall project PIs (Drs. Egan, Yaghjyan, and Goel) and other members of the research team. The committee will consider scientific merit, ethical and legal factors, and any potential conflicts of interest on the part of the applicant, with fair and transparent deliberation. Upon acceptance, de-identified samples and/or data will be released with Committee approval, and after establishing a Data Sharing or Material Transfer Agreement between institutions. The investigators have not budgeted costs for distribution of resources to end users. Hence, costs for data downloads or retrieval/ shipment of samples will be borne by researchers with approved protocols making the request. Further information can be obtained from the corresponding author.

## Results

By study close, we prospectively recruited 733 women in our biobanking study (348 from MCC, 153 from UF, and 232 from UM). The average age of women was 60 (range 40-92), with majority of the women (68.3%) being postmenopausal. This diverse sample included 49.2% Caucasian, 21.0% African American, 25.6% Hispanic, and 4.1% women from mixed or other races. Risk factor data collection was complete for 96.9% of women, with 57.8% and 64.8% of women also completing the optional diet and residential history questionnaire, respectively. Urine and saliva samples as well as mammograms were available for all women and stool samples along with the accompanying questionnaire were returned by 83% of participants.

Distribution of BI-RADS breast density categories from mammography reports in our sample (15.3% for BI-RADS A [almost entirely fatty], 46.7% for BI-RADS B [scattered areas of fibroglandular density], 34.4% for BI-RADS C [heterogeneously dense], and 3.7% for BI-RADS D [extremely dense]). Table 1 describes distribution of some of the breast cancer risk factors in our study population, overall, by study site, and by race/ethnicity.

**Table 1.** Characteristics of the study population, by study site and race/ethnicity^1^.

| Characteristic | Overall | MCC | UF | UM | White | African American | Hispanic | Other |
| --- | --- | --- | --- | --- | --- | --- | --- | --- |
| **Mean** |  |  |  |  |  |  |  |  |
| Age (years) | 59.68 | 61.00 | 57.67 | 59.04 | 61.68 | 59.42 | 56.92 | 54.34 |
| Age at menarche (years) | 12.41 | 12.48 | 12.28 | 12.39 | 12.52 | 12.34 | 12.27 | 12.30 |
| Body Mass Index (kg/m<sup>2</sup>) | 29.27 | 28.27 | 31.79 | 29.09 | 28.21 | 32.70 | 28.95 | 26.36 |
| Age at menopause (years) | 47.33 | 47.31 | 44.75 | 49.00 | 48.22 | 43.82 | 48.18 | 49.58 |
| Alcohol, drinks/week | 2.69 | 2.96 | 2.80 | 2.22 | 3.32 | 1.34 | 2.31 | 4.58 |
| **Percentages** |  |  |  |  |  |  |  |  |
| Parity/age at first birth |  |  |  |  |  |  |  |  |
| Nulliparous | 23.02 | 24.04 | 19.18 | 24.00 | 25.43 | 24.16 | 18.23 | 17.86 |
| Parous, age<25 years | 34.46 | 29.97 | 52.05 | 29.33 | 26.86 | 56.38 | 33.70 | 17.86 |
| Parous, age≥25 years | 42.51 | 45.99 | 28.08 | 46.67 | 47.71 | 19.46 | 48.07 | 64.29 |
| Family history of breast cancer (yes) | 23.52 | 30.06 | 19.59 | 16.37 | 28.65 | 22.15 | 16.30 | 14.29 |
| History of benign breast disease (yes) | 40.99 | 52.08 | 30.97 | 31.08 | 47.28 | 28.19 | 40.22 | 35.71 |
| Smoking |  |  |  |  |  |  |  |  |
| Never smoked | 67.84 | 70.15 | 59.46 | 69.91 | 62.93 | 77.18 | 67.39 | 82.14 |
| Past smoker | 25.25 | 26.87 | 24.32 | 23.45 | 31.32 | 14.77 | 24.46 | 10.71 |
| Current smoker | 6.91 | 2.99 | 16.22 | 6.64 | 5.75 | 8.05 | 8.15 | 7.14 |
| Menopausal status/MHT use |  |  |  |  |  |  |  |  |
| Premenopausal | 31.73 | 28.19 | 33.11 | 36.16 | 26.22 | 29.14 | 40.44 | 57.14 |
| Postmenopausal/never used MHT | 46.12 | 42.73 | 50.00 | 48.66 | 41.79 | 56.29 | 46.99 | 39.29 |
| Postmenopausal/past MHT | 12.98 | 18.40 | 6.08 | 9.38 | 18.16 | 8.61 | 8.20 | 3.57 |
| Postmenopausal/current MHT | 9.17 | 10.68 | 10.81 | 5.80 | 13.83 | 5.96 | 4.37 | 0.00 |
| BI-RADS breast density |  |  |  |  |  |  |  |  |
| Almost entirely fat (A) | 15.27 | 11.49 | 14.97 | 21.12 | 13.61 | 20.53 | 16.04 | 3.45 |
| Scattered areas of fibroglandular density (B) | 46.63 | 43.10 | 50.34 | 49.57 | 45.00 | 56.29 | 44.39 | 31.03 |
| Heterogeneously dense (C) | 34.39 | 41.67 | 32.65 | 24.57 | 37.78 | 23.17 | 32.62 | 62.07 |
| Extremely dense (D) | 3.71 | 3.74 | 2.04 | 4.74 | 3.61 | 0.00 | 6.95 | 3.45 |

Abbreviations: MCC – Moffitt Cancer Center; UF – University of Florida; UM – University of Miami
<sup>1</sup>Among 733 participants, the results reflect the numbers based on completed questionnaire data

## Discussion

As described above, over an approximately 3-year period we successfully enrolled 733 women with breast cancer risk factor data, urine, stool, and saliva samples, and mammogram collection. Of note, funding of the project coincided with emergence of the Covid19 pandemic such that recruitment was low during the initial project year and gradually increased to the originally projected levels of 8-10 participants per week as clinic operations returned to normal. Enrollment continued at the three project sites through June 2024 with a final sample size of 733 participants. This valuable resource offers unique opportunities supporting the study of the gut microbiome and breast cancer etiology.

Our imaging center-based recruitment efforts have generated a sample whose demographics mirror those of the state of Florida. We have been particularly successful in recruiting African American and Hispanic women who have been traditionally less likely to participate in research. We recognize, however, several challenges in covering broader recruitment areas and engaging additional imaging centers that could enable us to increase the cohort size. Thus, we are now working with additional facilities in the state to identify appropriate supplemental recruitment venues to further increase representation of African American and other underrepresented women in the study.

There are several advantages to our recruitment and data/sample collection procedures. The study consent, and most of the data collection, and urine/saliva collection were completed in the clinic thus allowing us to achieve collection of urine samples from all enrolled women as well as risk factor survey data from the great majority (97%) of participants. Collection of saliva for future genotyping studies is less invasive and time consuming than blood sample collection, thus improving participation. While stool sample collection is challenging and requires more commitment on the part of the participant, we were able to achieve a relatively high rate of stool sample collection (∼83%). Incorporation of $20 incentive payment for stool return likely encouraged participation.

Recruitment processes aimed to minimize each woman’s time commitment while maximizing the amount of collected information. Women who did not have sufficient time to complete survey in the clinic, were offered an opportunity to do so at home or were followed up by the study coordinators who offered coordinator-assisted survey completion by phone, if preferred. We believe these approaches allowed us to maximize the amount of collected information and ensured high rates of completion.

In the initial months of sample collection using a USPS sample return delivery protocol, shipment times lagged especially for UF and UM. Therefore, we adopted FedEx-assisted delivery which reduced return times by several days on average. Previous studies show reasonable levels of stability of the gut microbiome under field conditions even with large variation in timing of collection and fluctuation of temperatures prior to freezer storage [41]. Urine samples were aliquoted and stored at −80ºC at each institution within variable amount of time (on average 55 minutes for MCC, 49 minutes for UF, and 2 hours for UM). Our previous pilot investigation with a similar recruitment protocol for a study of gut microbiome and estrogens has demonstrated stable concentrations of estrogen metabolites with varying collection-to-storage times.\

## Conclusions

We established a cohort of screening-aged women with collection of breast cancer risk factor data, urine, stool, and saliva samples, and mammographic breast density in the State of Florida. By using this unique resource, investigators have the opportunity to explore lifestyle factors as well as circulating biomarkers in relation to various health end-points in women and to examine contributions of gut microbiome to breast health via related intermediate phenotypes, such as circulating estrogens and mammographic breast density. Investigators encourage collaboration with cancer researchers in Florida and beyond.

## Conflict of interest

The authors declare that they have no conflict of interest.

## Funding Statement

This project was funded by the Florida Department of Health Bankhead-Coley Cancer Research Program Infrastructure Grant.

## Acknowledgements

This work has been supported in part by the Participant Research, Interventions, and Measurement Core and the Tissue Core Facilities at the H. Lee Moffitt Cancer Center & Research Institute, an NCI designated Comprehensive Cancer Center (P30-CA076292).

## Notes

### Competing Interest Statement

The authors have declared no competing interest.

### Author Declarations

Advarra Institutional Review Board of Moffitt Cancer Center gave ethical approval for this work. Health Center Institutional Review Board of the University of Florida gave ethical approval for this work. Institutional Review Board of the University of Miami gave ethical approval for this work.

## References

1. Jemal A, Ward EM, Johnson CJ, Cronin KA, Ma J, Ryerson B, et al. Annual Report to the Nation on the Status of Cancer, 1975-2014, Featuring Survival. Journal of the National Cancer Institute 2017;109(9)

2. Fraune S, Bosch TC. Why bacteria matter in animal development and evolution. Bioessays 2010;32(7):571–80

3. Ley RE, Peterson DA, Gordon JI. Ecological and evolutionary forces shaping microbial diversity in the human intestine. Cell 2006;124(4):837–48

4. Cani PD. Human gut microbiome: hopes, threats and promises. Gut 2018;67(9):1716–25

5. Zeevi D, Korem T, Godneva A, Bar N, Kurilshikov A, Lotan-Pompan M, et al. Structural variation in the gut microbiome associates with host health. Nature 2019;568(7750):43–8

6. Sekirov I, Russell SL, Antunes LC, Finlay BB. Gut microbiota in health and disease. Physiol Rev 2010;90(3):859–904

7. Castaner O, Goday A, Park YM, Lee SH, Magkos F, Shiow STE, et al. The Gut Microbiome Profile in Obesity: A Systematic Review. International journal of endocrinology 2018;2018:4095789

8. Tseng CH, Wu CY. The gut microbiome in obesity. Journal of the Formosan Medical Association = Taiwan yi zhi 2019;118 Suppl 1:S3–s9

9. Maruvada P, Leone V, Kaplan LM, Chang EB. The Human Microbiome and Obesity: Moving beyond Associations. Cell host & microbe 2017;22(5):589–99

10. Zhu J, Liao M, Yao Z, Liang W, Li Q, Liu J, et al. Breast cancer in postmenopausal women is associated with an altered gut metagenome. Microbiome 2018;6(1):136

11. Tilg H, Kaser A. Gut microbiome, obesity, and metabolic dysfunction. The Journal of clinical investigation 2011;121(6):2126–32

12. Kinross JM, Darzi AW, Nicholson JK. Gut microbiome-host interactions in health and disease. Genome medicine 2011;3(3):14

13. Fernandez MF, Reina-Perez I, Astorga JM, Rodriguez-Carrillo A, Plaza-Diaz J, Fontana L. Breast Cancer and Its Relationship with the Microbiota. International journal of environmental research and public health 2018;15(8)

14. Miko E, Kovacs T, Sebo E, Toth J, Csonka T, Ujlaki G, et al. Microbiome-Microbial Metabolome-Cancer Cell Interactions in Breast Cancer-Familiar, but Unexplored. Cells 2019;8(4)

15. Schwabe RF, Jobin C. The microbiome and cancer. Nature reviews Cancer 2013;13(11):800–12

16. Kwa M, Plottel CS, Blaser MJ, Adams S. The Intestinal Microbiome and Estrogen Receptor-Positive Female Breast Cancer. Journal of the National Cancer Institute 2016;108(8)

17. Baker JM, Al-Nakkash L, Herbst-Kralovetz MM. Estrogen-gut microbiome axis: Physiological and clinical implications. Maturitas 2017;103:45–53

18. Zmora N, Soffer E, Elinav E. Transforming medicine with the microbiome. Science translational medicine 2019;11(477)

19. Horwitz RI, Feinstein AR. The problem of “protopathic bias” in case-control studies. Am J Med 1980;68(2):255–8

20. Carey RM. New Intermediate Phenotype of Resistant Hypertension. Hypertension 2017;69(5):789–90

21. Thomas DC, Conti DV, Baurley J, Nijhout F, Reed M, Ulrich CM. Use of pathway information in molecular epidemiology. Human Genomics 2009;4(1):21

22. Windle M. Intermediate phenotypes for alcohol use and alcohol dependence: Empirical findings and conceptual issues. Genetic influences on addiction: An intermediate phenotype approach. Cambridge, MA, US: Boston Review; 2013. p 257–74.

23. Camilleri M, Katzka DA. Irritable Bowel Syndrome: Methods, Mechanisms, and Pathophysiology. Genetic epidemiology and pharmacogenetics in irritable bowel syndrome. American Journal of Physiology-Gastrointestinal and Liver Physiology 2012;302(10):G1075-G84

24. Sudenga SL, Shrestha S. Key considerations and current perspectives of epidemiological studies on human papillomavirus persistence, the intermediate phenotype to cervical cancer. International Journal of Infectious Diseases 2013;17(4):e216-e20

25. Subar AF, Thompson FE, Kipnis V, Midthune D, Hurwitz P, McNutt S, et al. Comparative validation of the Block, Willett, and National Cancer Institute food frequency questionnaires: the Eating at America’s Table Study. Am J Epidemiol 2001;154(12):1089–99

26. Thompson FE, Subar AF, Brown CC, Smith AF, Sharbaugh CO, Jobe JB, et al. Cognitive research enhances accuracy of food frequency questionnaire reports: results of an experimental validation study. Journal of the American Dietetic Association 2002;102(2):212–25

27. Goel N, Hernandez AE, Antoni MH, Kesmodel S, Pinheiro PS, Kobetz E, et al. ZIP Code to Genomic Code: Neighborhood Disadvantage, Aggressive Breast Cancer Biology, and Breast Cancer Outcomes. Ann Surg 2024;280(1):1–10

28. Yaghjyan L, Mai V, Wang X, Ukhanova M, Tagliamonte M, Martinez YC, et al. Gut microbiome, body weight, and mammographic breast density in healthy postmenopausal women. Cancer Causes Control 2021;32(7):681–92

29. Yaghjyan L, Mai V, Darville LNF, Cline J, Wang X, Ukhanova M, et al. Associations of gut microbiome with endogenous estrogen levels in healthy postmenopausal women. Cancer Causes Control 2023;34(10):873–81

30. Heine JJ, Scott CG, Sellers TA, Brandt KR, Serie DJ, Wu FF, et al. A novel automated mammographic density measure and breast cancer risk. J Natl Cancer Inst 2012;104(13):1028–37

31. Vachon CM, Fowler EE, Tiffenberg G, Scott CG, Pankratz VS, Sellers TA, et al. Comparison of percent density from raw and processed full-field digital mammography data. Breast Cancer Res 2013;15(1):R1

32. Heine JJ, Cao K, Rollison DE, Tiffenberg G, Thomas JA. A quantitative description of the percentage of breast density measurement using full-field digital mammography. Acad Radiol 2011;18(5):556–64

33. Vachon CM, Fowler EE, Tiffenberg G, Scott CG, Pankratz VS, Sellers TA, et al. Comparison of percent density from raw and processed full-field digital mammography data. Breast Cancer Res 2013;15(1):R1

34. Heine JJ, Carston MJ, Scott CG, Brandt KR, Wu FF, Pankratz VS, et al. An automated approach for estimation of breast density. Cancer Epidemiol Biomarkers Prev 2008;17(11):3090–7

35. Eng A, Gallant Z, Shepherd J, McCormack V, Li J, Dowsett M, et al. Digital mammographic density and breast cancer risk: a case-control study of six alternative density assessment methods. Breast cancer research: BCR 2014;16(5):439

36. Boyd NF, Martin LJ, Yaffe MJ, Minkin S. Mammographic density and breast cancer risk: current understanding and future prospects. Breast cancer research: BCR 2011;13(6):223

37. Harvey JA, Bovbjerg VE. Quantitative assessment of mammographic breast density: relationship with breast cancer risk. Radiology 2004;230(1):29–41

38. McCormack VA, dos Santos Silva I. Breast density and parenchymal patterns as markers of breast cancer risk: a meta-analysis. Cancer Epidemiol Biomarkers Prev 2006;15(6):1159–69

39. Pettersson A, Graff RE, Ursin G, Santos Silva ID, McCormack V, Baglietto L, et al. Mammographic density phenotypes and risk of breast cancer: a meta-analysis. J Natl Cancer Inst 2014;106(5)

40. Byng JW, Boyd NF, Little L, Lockwood G, Fishell E, Jong RA, et al. Symmetry of projection in the quantitative analysis of mammographic images. Eur J Cancer Prev 1996;5(5):319–27

41. Song SJ, Amir A, Metcalf JL, Amato KR, Xu ZZ, Humphrey G, et al. Preservation Methods Differ in Fecal Microbiome Stability, Affecting Suitability for Field Studies. mSystems 2016;1(3)

